# Use it or lose it: A four-year follow-up to assess whether engagement with physical activity close to one’s physical capacity may protect against decline in physical functioning among older adults

**DOI:** 10.1101/2024.09.27.24314462

**Authors:** Antti Löppönen, Katja Lindeman, Lotta Palmberg, Evelien Van Roie, Christophe Delecluse, Erja Portegijs, Taina Rantanen, Timo Rantalainen, Laura Karavirta

## Abstract

**PURPOSE:** Physical activity (PA) is distinct from physical capacity (PC), even though they are correlated in old age. PC defines the limits for PA, while activities in daily life typically remain submaximal. Individuals whose intensity of daily activities is close to physical capacity may be better protected from future decline in physical function compared to those who do not, although prospective research to support this hypothesis is lacking. Therefore, this study compared changes in physical function over a four-year follow-up between community-dwelling older adults categorized based on their combined baseline PC and PA.

**METHODS:** This was a four-year longitudinal follow-up study of older adults aged 75-85 years at baseline (N = 312, 60% women). Baseline PC was determined based on 5-second Mean Amplitude Deviation (MAD) epoch value during the maximal 10-meter walking test, and PA was determined based on the peak 75-minutes MAD intensity threshold from thigh-worn accelerometer monitoring over 3-7 days. Baseline values of PA and PC were categorized into lowPC-lowPA, lowPC-highPA, highPC-lowPA, and highPC-highPA profiles. Physical function was evaluated using the Short Physical Performance Battery (SPPB) at baseline and at the follow-up, with total score and 5 x Sit-To-Stand (5xSTS) test time as the primary outcomes. Nonparametric tests and generalized estimating equations were used for analyses.

**RESULTS:** From baseline to follow-up, statistically significant changes in the SPPB total score and 5xSTS test time were observed in all profiles (p<0.05) except the low PC-high PA profile. Over the follow-up period, the decrease was greater for low versus high PA profiles within both PC profiles for SPPB total score (high PC: B -0.61, SE 0.24, 95% CI -1.08, -0.15; low PC: B -0.96, SE 0.35, 95% CI -1.62, -0.32), but not for 5xSTS time. No statistically significant difference was observed in the change in 5xSTS test time between the low and high PA profiles for either PC profile.

**CONCLUSIONS:** The findings suggest that engaging in demanding PA regardless of baseline PC may help to protect against a decline in physical functioning in old age. Consequently, older adults should be encouraged to engage in physically demanding activities that could potentially enhance their functional capacity.

## INTRODUCTION

Physical functioning has been identified as an important factor that enables independent living among older adults [1]. The most effective way to maintain physical functioning is through diverse and sufficiently challenging physical activity and exercise, which should also include activities tailored to the individual’s capabilities [2]. However, multiple internal and external factors, such as ability to walk and social support, affect physical behaviour resulting in considerable variation in daily physical activity between older adults [3,4].

Physical activity has been presented as a distinct construct from physical capacity [5–11]. Although physical capacity correlates with physical activity, both entail a distinct construct. Physical capacity primarily defines the limits of physical activity rather than ensuring that individuals use their full capacity in daily life [10,11]. Based on locomotory system mechanobiology (i.e., disuse causes locomotory system atrophy, increased use hypertrophy [12]) one would expect functional capacity to be lost if full capacity is seldom utilised, i.e., the use it or lose it -principle. This demarcation between activity and capacity and the associated use-dependent adaptations have been operationalized in the physical capacity-physical activity (PC-PA) concept proposed by Koolen et al. and Orme et al. which categorizes individuals into four profiles: low PC -low PA (“cannot do – does not do”), low PC -high PA (“cannot do, does do”), high PC -low PA (“can do, does not do”) and high PC -high PA (“can do, does do”) [13,14].

To our knowledge, no studies have focused on PC-PA profiles in older adults in a longitudinal setting, which allows prediction of future conditions based on the profiles. Additionally, the methods used to determine PC and PA have not been “apples-to-apples” comparisons. Rather, the operationalization has been, e.g., PC determined by walking distance or the Timed Up and Go (TUG) compared with PA estimated by daily step counts [13,15]. Orme and colleagues introduced in their technical note that PC-PA profiles can be created by instrumenting a walking test and free-living PA with an accelerometer [14], thereby ensuring a direct “apples-to-apples” comparison of the intensity of PC and PA using acceleration. We propose addressing PA and PC intensities from free-living and standardized testing, respectively, using 5-second Mean Amplitude Deviation (MAD) epochs [16]. This approach allows for the examination of the PA intensity distribution, which can be used to determine the vigorous MAD intensity threshold -the MAD value that corresponds to the 75 minutes per week of physical activity recommended by the World Health Organization (WHO) [17]. As such, it is well-suited for defining ‘does do, does not do’ in PA. This is meaningfully comparable to PC, which was defined as the MAD during a 10-meter walking test [18].

So far, there are no prospective studies shedding light on whether older people who approach their capacity in free-living activities are better protected against future decline in physical function. Therefore, the aim of this study was to compare changes in physical function over four years between community-dwelling older adults categorized on the basis of their baseline PC and PA. We hypothesized that those older adults who were physically active close to their physical capacity would experience a slower decline in physical function.

## METHODS

### Participants and design

The data for this observational study were drawn from data collected in the AGNES study (Active Aging -Resilience and external support as modifiers of the disablement outcome; n = 1 021), which was conducted at the Gerontology Research Center, University of Jyväskylä [19]. The AGNES study comprises three age cohorts (75, 80, and 85 years of age) of people living independently in the city of Jyväskylä, in Central Finland. The baseline data were collected in 2017-2018 and the 4-year follow-up measurements were carried out in 2021-2022. The Ethical Committee of the Central Finland Health Care District provided an ethical statement on the research plan and protocol of the AGNES baseline (August 23, 2017) and follow-up study (September 8, 2021). The study was executed in accordance with the principles of the Declaration of Helsinki and all participants gave written informed consent.

Baseline recruitment was drawn as a random sample from postcode areas in Jyväskylä, Finland, using the registers of the Digital and Population Data Services Agency in Finland. Baseline inclusion criteria were age and residence in the study area, willingness to participate and the ability to communicate [19]. After exclusions, 1021 individuals participated in the study, of whom 432 wore a tri-axial accelerometer for 3 to 7 consecutive days and participated in the 10-meter walking test with an accelerometer, forming a baseline sample for this study. Of the older adults included in the baseline sample, 26 were deceased, 6 could not be reached, 77 were not interested or not applicable, and 11 had not completed the Short Physical Performance Battery (SPPB) at the time of the follow-up, resulting in a final follow-up sample of 312 participants. All of these participants had at least 3 days of successful accelerometer recording and completed the maximal 10-meter walking test at baseline, and also participated in the 4-year follow-up home interview with the SPPB test.

### Determination of PC-PA profiles

At baseline, a research assistant visited the participant’s home, conducted a face-to-face interview, and placed an accelerometer on the participant’s thigh using a waterproof film. Participants wore the accelerometer continuously (24 hours a day) for 3 to 7 consecutive days (Figure 1). Subsequently, participants arrived in the laboratory and participated in a comprehensive health and physical function assessment protocol, which included a maximal 10-meter walking test.

**Figure 1.**
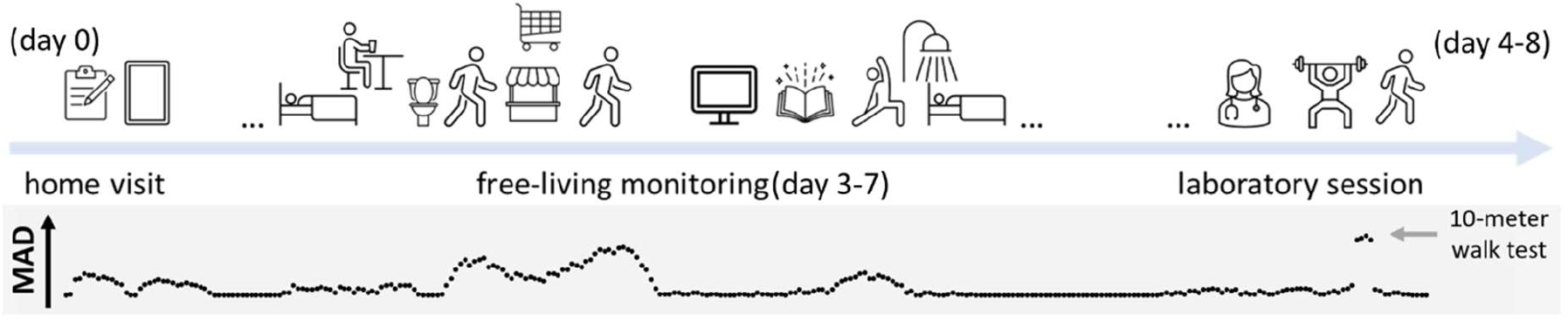
Timeline of study protocol and typical occurrence of 5-second MAD epochs.

The study employed a UKK RM42 tri-axial accelerometer (13-bit analog-to-digital conversion, acceleration range ±16 g, UKK Terveyspalvelut Oy, Tampere, Finland). The accelerometer sampling rate was set at 100 samples per second, with acceleration recorded in gravity units. From the data collected by the accelerometer, the mean amplitude deviation (MAD = 1/n *∑|rk –r|) of each 24-hour period was calculated based on the vector magnitude (Euclidian norm) of the resultant acceleration (√x2+y2+z2) in non-overlapping 5-second epochs, following the methodology outlined in previous reports [16,20].

#### Laboratory-based assessment of physical capacity (PC) and determination of ‘Can do’ and ‘Cannot do’ groups

Physical capacity (PC) was determined based on the mean amplitude deviation (MAD) epoch during the maximal 10-meter walking test [21,22] (Figure 2A). The test was conducted in a research laboratory walking track, with a total distance of 20 meters reserved for the test. The test area consisted of a 5-meter acceleration phase, a 10-meter test distance and a 5-meter deceleration phase. During the maximal 10-meter walking test, participants were instructed to walk safely from the starting point to the end point at their maximum walking speed. The MAD value during the test was manually extracted from the data by visually following the test protocol in the data. The MAD value during 10-meter walking test was strongly correlated with maximum walking speed of same test in this dataset (n = 432, r = 0.72, p < .001).

**Figure 2.**
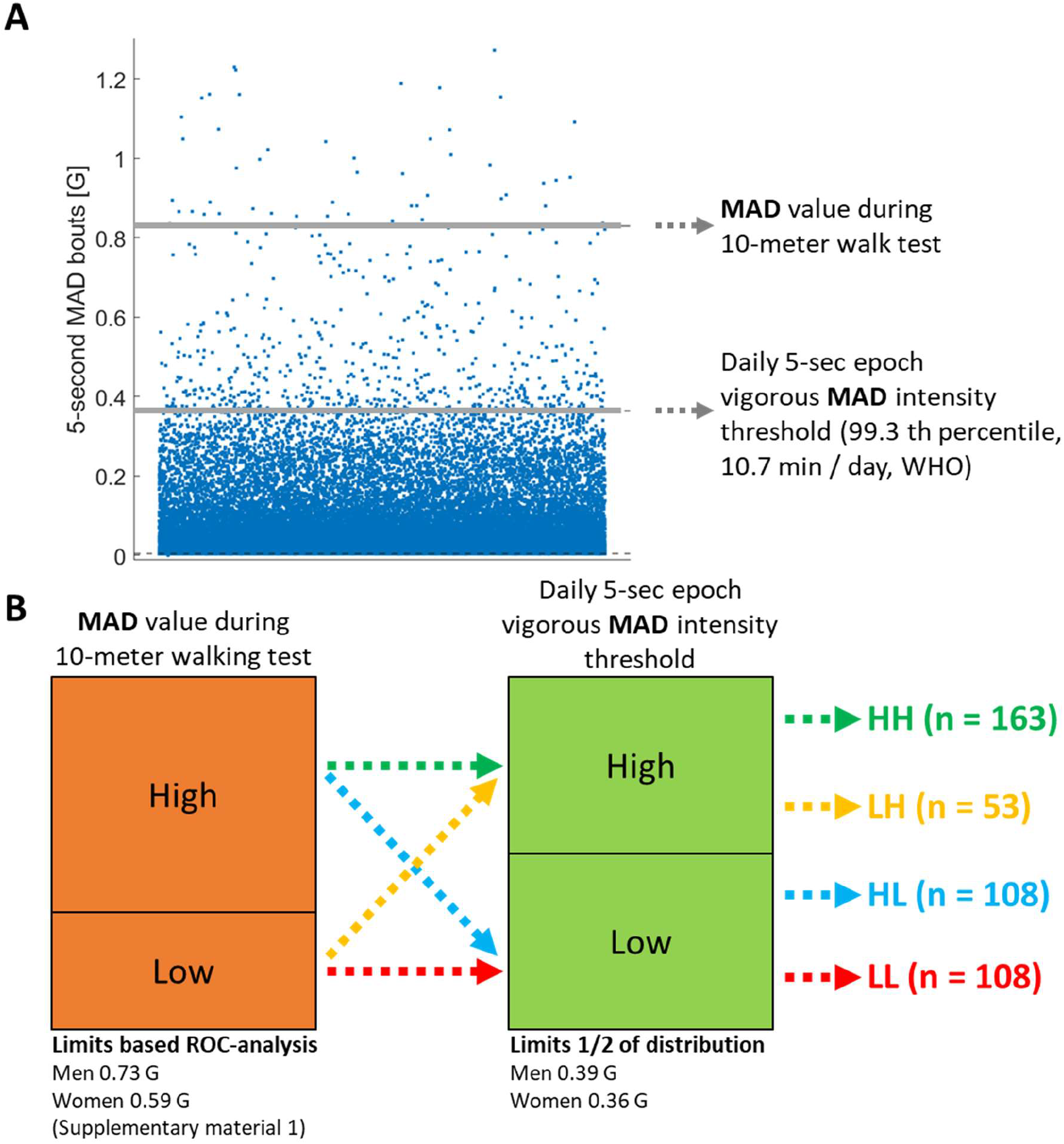
A. Specification of the variables used for PC-PA profiling. **B**. Generating PC-PA profiles using the MAD value during the 10-meter walking test and the peak 75-minutes MAD intensity threshold.

To the best of the authors’ knowledge, there are no thresholds for the MAD value during the maximal 10-meter walking test for this age group (75-85 years), based on which capacity can be classified as non-limited and limited physical functioning [23]. Therefore, in this study, the thresholds for the MAD value during 10-meter walking test were determined separately for men and women using Receiver Operating Characteristics (ROC) in classifying the dataset into high (SPPB ≥ 10) and low (SPPB < 10) physical functioning [24]. For men, a cut-point of 0.73 G was determined with moderate accuracy (specificity 70%, sensitivity 66%, AUC 0.76), and similarly, for women, the cut-point was 0.59 G (specificity 68%, sensitivity 76%, AUC 0.73). More detailed analysis can be found in Supplementary File 1.

#### Physical activity (PA) assessment and the determination of ‘Does do’ and ‘Does not do’ groups

The ‘Does do’ and ‘Does not do’ groups were defined throughout the entire free-living monitoring period (aggregating full 24-hour measurement days) using the 5-second MAD epoch distribution (intensity profile), which describes the dispersion of activity intensities. The highest part of the distribution represents the highest attained intensity [25] (Figure 2A). The peak 75-min MAD intensity threshold of the 5-second MAD epoch distribution was determined to represent the threshold for 75 minutes per week (10.7 minutes per day). This threshold represents the 99.3rd percentile of the activity intensity distribution. It indicates that the activity intensity exceeds this level only 0.7% of the time each day, which equates to 10.7 minutes out of the total 1440 minutes in a day. The ‘Does do’ and ‘Does not do’ groups were defined via a data-driven approach by dividing the vigorous MAD intensity threshold into two equally sized groups separately for men and women (Figure 2B).

#### Generation of PC-PA profiles

Finally, the PC and PA categories were combined to create four profiles: LL = low PC – low PA (“cannot do – does not do”); LH = low PC – high PA (“cannot do, does do”); HL = high PC – low PA (“can do, does not do”); and HH = high PC – high PA (“can do, does do”) following the approach presented by Koolen et al. and Orme et al. [13,14] (Figure 2B).

### Lower extremity functioning as a follow-up outcome

For the longitudinal analyses, lower extremity functioning was assessed at baseline and follow-up in the participants’ homes using the Short Physical Performance Battery (SPPB). The SPPB comprised tests of standing balance, walking speed over a 3-meter distance, and the five-times-sit-to-stand (5xSTS) test [26,27]. In this study, we used the SPPB total score (maximum of 12 points, with higher scores indicating better lower extremity functioning) and the time in seconds of the 5xSTS test as outcomes.

### Descriptive Characteristics and Other Measurements

Age and sex were obtained from the population register and cognitive function was assessed using standardized procedures (Mini-Mental State Examination, MMSE [28]).

*Walking difficulties* over distances of 500 meters and 2 kilometres were investigated by asking the participants, “Do you have difficulty walking 2 kilometres / 500 meters?” The response options included: 1) able to manage without difficulty, 2) able to manage with some difficulty, 3) able to manage with a great deal of difficulty, 4) able to manage only with the help of another person, and 5) unable to manage even with help. In this study, response options 2-5 were grouped into the category “I have walking difficulties,” with response option 1 indicating “I do not have walking difficulties” [29,30].

*Self-reported health status* was assessed with the question: “How would you rate your current overall health?” The response options were: “1. excellent”, “2. good”, “3. fair”, “4. poor”, “5. very poor”. For the analysis, response options 1 and 2 were categorized as “good perceived health” and response options 3-5 were combined into the category “limited perceived health”. *The perceived ability to perform desired activities* from the perspective of health was assessed with the question: “To what extent has your health or physical ability prevented you from doing the things you wanted to do in the past 4 weeks?” The response options were: “1. not at all”, “2. a little”, “3. somewhat”, “4. a lot”, “5. extremely.” For the analysis, response option 1 was categorized as “my health does not prevent me from doing things I want” and response options 2-5 were combined into the category “my health prevents me from doing things I want” [31].

*Maximal isometric knee extension strength* was assessed in the laboratory (at a knee angle of 60 degrees from the fully extended leg to flexion) of the dominant leg in a seated position using an adjustable dynamometer chair (Metitur LTD, Jyväskylä, Finland). At least three attempts were required, and the highest force (N) was chosen for the analysis [32]. *Maximal isometric handgrip strength* was measured on the dominant side with a hand-held adjustable dynamometer (Jamar Plus digital hand dynamometer, Patterson Medical, 6 Cedarburg, WI, USA) and expressed in kg [33].

### Statistical analyses

Baseline comparisons between the different profiles were made across the entire sample of 432 older adults. Baseline descriptive data are presented as means and standard deviations (SD) for continuous variables and relative frequencies (%) for dichotomous variables. Differences in baseline characteristics between PC-PA profiles were tested by independent samples Kruskal-Wallis test for continuous variables and by Chi-square test for dichotomous variables. Pairwise comparisons between profiles were Bonferroni-corrected.

Changes over four years in SPPB scores and 5xSTS test time were examined for a longitudinal sample (n = 312) who had data from both baseline and follow-up measurements. The Wilcoxon signed rank test for related samples was used to analyze changes within the different PC-PA profiles in the total SPPB score and in the 5xSTS test time. Generalized estimating equations (GEE) [34] with a linear link function and unstructured working correlation matrix were used to determine whether the SPPB total score and the 5xSTS test time (group effect) and their change over time (group by time interaction) differed between low and high PA profiles. Analyses were performed separately for low PC and high PC profiles. All GEE models were adjusted for sex, age cohort, perceived health status, days included in the accelerometry analysis and 10m MAD. Population-averaged coefficients (B), standard errors (SE), and 95% confidence intervals are reported. Statistical significance was set at p < 0.05, and statistical analyses were performed using the SPSS statistical software package (IBM Corp. Released 2021. IBM SPSS Statistics for Windows, version 28.0. Armonk, NY: IBM Corp.) [35]. Figures were generated in the “R” statistical environment (version 4.3.1) [36].

## RESULTS

### Baseline characteristics of the generated PC-PA profiles

The descriptive baseline characteristics of the PC-PA profiles for the entire sample of 432 older adults are presented in Table 1. According to the baseline characteristics, the low PC profiles differed from each other in terms of 10-meter walking speed, SPPB total score, self-reported walking difficulty at 500 m and 2 km distances, self-reported health status and self-reported limited ability to perform desired activities. The high PC profiles differed from each other in terms of age and self-reported walking difficulty in 500 m and 2 km distances (Table 1).

**Table 1.** Baseline characteristics according to the PC-PA profiles (n = 432)

| Group | LL<br>low PC-low PA<br>($n = 108$) | LH<br>low PC-high PA<br>($n = 53$) | HL<br>high PC-low PA<br>($n = 108$) | HH<br>high PC-high PA<br>($n = 163$) | p-value |
| --- | --- | --- | --- | --- | --- |
| Women | 58 % | 49 % | 61 % | 63 % | .321 ^ |
| Age | 79.1 (3.9) | 77.6 (3.2) | 78.9 (3.3) | 77.6 (3.1) | .001 * LL-HH, HL-HH |
| MMSE | 27.2 (2.6) | 26.9 (2.6) | 27.4 (2.2) | 28.0 (1.9) | .005 * LH-HH |
| Leg strength [N] | 302 (109) | 345 (110) | 348 (108) | 372 (110) | <.001 * LL-HL, LL-HH |
| Grip strenght [kg] | 30.3 (10.7) | 34.2 (12.4) | 32.4 (10.5) | 33.3 (10.7) | .076 * |
| 10-meter maximal walking speed [m/s] | 1.44 (0.29) | 1.75 (0.26) | 1.90 (0.33) | 2.00 (0.32) | <.001 * LL-LH, LL-HL, LL-HH, LH-HH |
| 5xSTS time [s] | 14.3 (4.3) | 12.8 (3.6) | 11.4 (3.2) | 11.7 (2.9) | <.001 * LL-HL, LL-HH |
| SPPB total score | 9.4 (2.1) | 10.2 (1.8) | 10.9 (1.3) | 11.0 (1.2) | <.001 * LL-LH, LL-HL, LL-HH |
| SR walk difficulty 500 m | 36 % | 9 % | 16 % | 3 % | <.001 ^ LL-LH, LL-HL, LL-HH, HL-HH |
| SR walk difficulty 2 km | 54 % | 15 % | 27 % | 7 % | <.001 ^ LL-LH, LL-HL, LL-HH |
| SR health status is weak | 71 % | 40 % | 42 % | 37 % | <.001 ^ LL-LH, LL-HL, LL-HH |
| SR limited ability to perform desired activities | 61 % | 25 % | 37 % | 23 % | <.001 ^ LL-LH, LL-HL, LL-HH |

Note. MMSE = Mini-Mental State Examination, STS = sit-to-stand transitions, SPPB= Short Physical Performance Battery, SR = self-reported/rated, ^ Chi-square test and pairwise comparison with Bonferroni correction, \* Independent-samples Kruskal-Wallis test and pairwise comparison with Bonferroni correction.

Figure 3 shows the baseline intensity distribution of PC-PA profiles together with their MAD values during the laboratory-based 10-meter maximal walking test (highlighted in orange) and the distribution of MAD values, including their vigorous MAD intensity threshold (shown in black), for free-living 5-second MAD epochs. According to the figure, the capacity of the HL (high PC-low PA) and HH (high PC-high PA) groups does not differ statistically (confidence intervals do not overlap), whereas the physical activity differs significantly between these groups.

**Figure 3.**
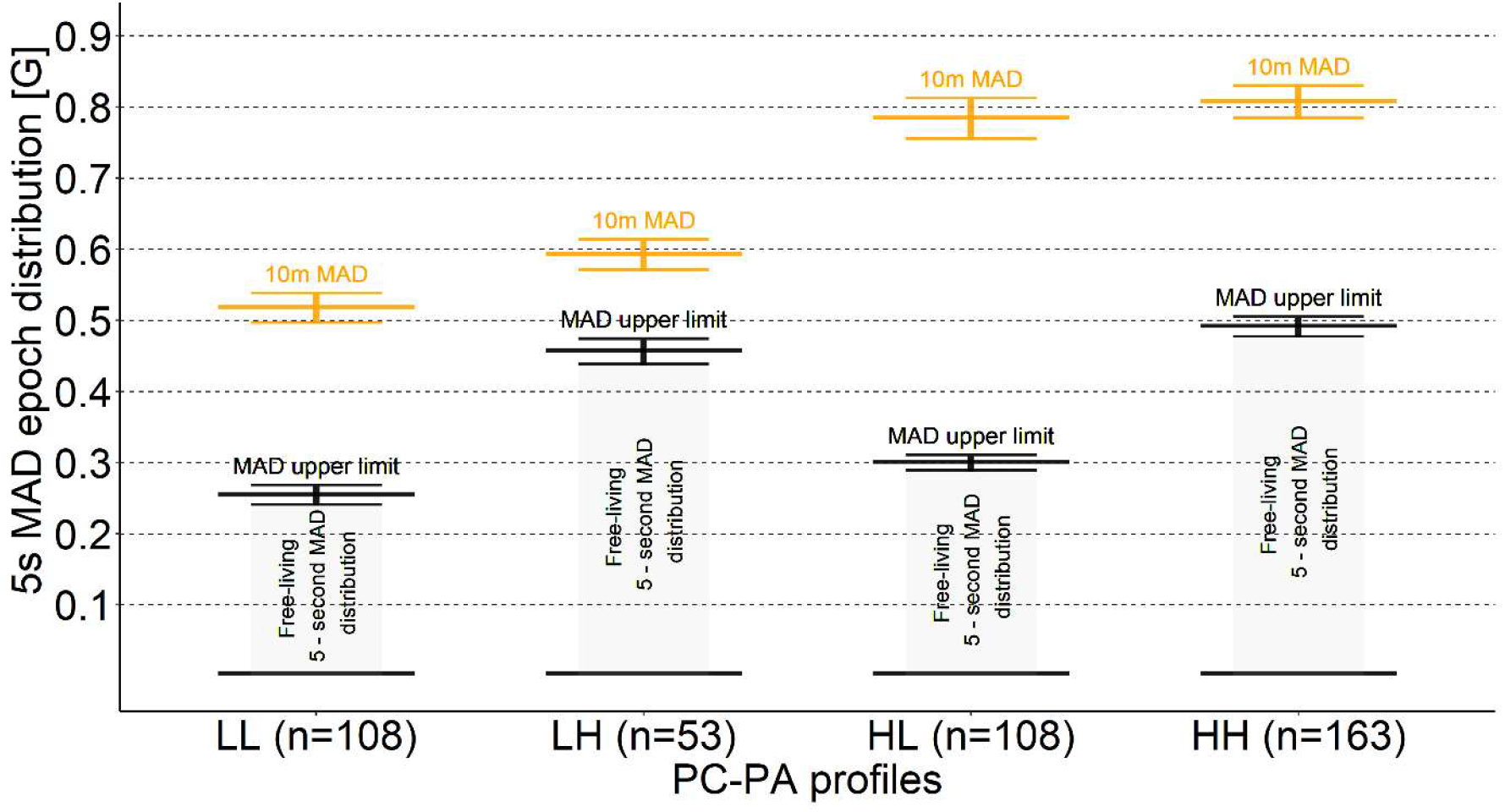
Distribution of the different PC-PA profiles according to the MAD value during the 10-meter walking test (highlighted in orange) and the peak 75-minutes MAD intensity threshold (99.3rd percentile) (shown in black).

### Changes in the SPPB total score and the 5xSTS time in a four-year follow-up

A longitudinal sample (n = 312) of those who participated in both baseline and follow-up measurements was used to examine the difference in change in the SPPB total score and the 5xSTS time within and between the different profiles over four years of follow-up. During follow-up, the total SPPB score decreased by at least 2 points in 43% of participants in the low PC -low PA profile, 18% of participants in the low PC -high PA profile, 29% of participants in the high PC -low PA profile, and 15% of participants in the high PC – high PA profile. The change in the SPPB total score and the 5xSTS test time within the different PC-PA profiles is shown in Table 2. From baseline to follow-up, statistically significant changes in SPPB total score and 5xSTS test time were observed in all other profile than in the low PC – high PA profile.

**Table 2.** Changes within the different PC-PA profiles in SPPB total score and 5xSTS test time over four years of follow-up (n = 312)

|  | **LL**<br>**low PC-**<br>**low PA**<br>**(n = 65)** | **LH**<br>**low PC-**<br>**high PA**<br>**(n = 39)** | **HL**<br>**high PC-**<br>**low PA**<br>**(n = 86)** | **HH**<br>**high PC-**<br>**high PA**<br>**(n = 122)** |
| --- | --- | --- | --- | --- |
|  | Mean (SD) | Mean (SD) | Mean (SD) | Mean (SD) |
| BL SPPB total score | 9.7 (2.0) | 10.5 (1.7) | 11.0 (1.3) | 10.9 (1.2) |
| FU SPPB total score | 8.3 (2.7) | 10.1 (1.7) | 10.1 (2.2) | 10.6 (1.5) |
| SPPB total score difference | -1.4 (2.0) | -0.4 (1.6) | -0.9 (1.9) | -0.3 (1.4) |
| *\*p-value* | **<.001** | .059 | **<.001** | **.013** |
|  | Mean (SD) | Mean (SD) | Mean (SD) | Mean (SD) |
| BL 5xSTS test time [s] | 13.6 (3.8) | 12.3 (3.3) | 11.2 (2.8) | 11.7 (3.0) |
| FU 5xSTS test time [s] | 15.7 (4.8) | 13.1 (3.3) | 12.8 (4.3) | 12.3 (3.4) |
| 5xSTS test time difference [s] | +2.0 (5.0) | +0.7 (3.2) | +1.6 (4.1) | +0.6 (3.2) |
| *\*p-value* | **.002** | .137 | **.005** | **.033** |

Note. BL = baseline 2017-2018; FU = follow-up 2021-2022; SPPB = Short Physical Performance Battery; 5xSTS = five-times-sit-to-stand test. \* Wilcoxon signed rank test for related samples

The results of the covariate-adjusted GEE models between high and low PA profiles, separately for low and high PC profiles, are presented in Table 3. Baseline level of the SPPB total score or 5xSTS test time did not differ between low and high PA profiles in either PC profile. Over the follow-up, the decrease in SPPB total score was greater for low PA profiles compared to high PA profiles in both PC profiles (high PC: p = 0.010, low PC: p = 0.006). For the 5xSTS test, the difference in change in test time between the low and high PA profiles did not reach statistical significance for either PC profile (high PC: p = 0.058, low PC: p = 0.107).

**Table 3.**
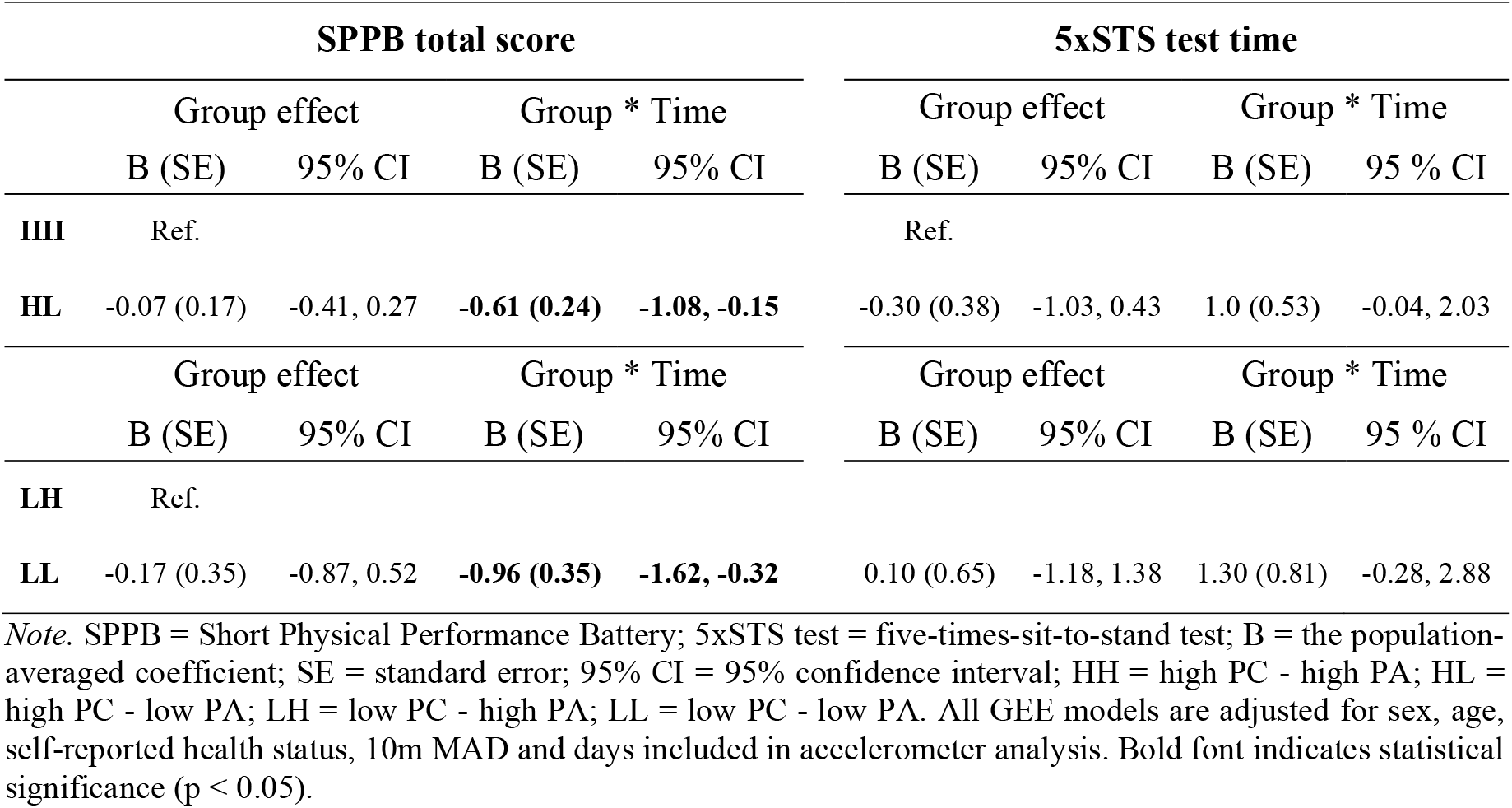
GEE model estimates for group effect and group-by-time interactions for SBBP total score and 5xSTS test time (n = 312).

## DISCUSSION

To increase our understanding of the effects of challenging one’s abilities for maintaining physical functioning in old age, this study compared changes in physical function over 4-year follow-up among community-dwelling older adults categorized based on their baseline physical capacity and physical activity. The changes observed from the baseline to the follow-up within the different profiles demonstrated a statistically significant change in the SPPB total score and 5xSTS test time in all profiles, with the exception of the low PC-high PA profile. Over the follow-up period, the decrease in the SPPB total score was deeper for low compared to high PA profiles in both PC profiles. However, no statistically significant difference was observed in the change in 5xSTS test time between the low and high PA profiles for either PC profile. Based on the changes within the profiles, our results suggest that ageing reduces lower extremity function in older adults regardless of their physical capacity or intensity of physical activity, but by challenging themselves to be active close to their capacity, it is possible to slow the decline in lower extremity function. Our results also suggest that older adults who already have limitations in physical function can maintain their level of functioning through physical activity.

In this study, the groups were formed based on the profiling presented by Koolen and colleagues [13]. The lower capacity groups showed statistically significant differences in terms of walking speed, 5xSTS test time, and maximum knee extension strength in the descriptive data. However, in the two higher capacity groups, no statistical differences were observed in functional ability and capacity variables. The descriptive profiles are consistent with the findings of Adams and colleagues [15], where the timed “up and go” (TUG) test, used as a basis for capacity assessment, showed differences among the lower capacity groups, unlike in the higher capacity groups. To better understand the decline in functional capacity from the perspective of the “use it or lose it” principle [37], this study focused particularly on the intensity profile of daily activities The 5-second MAD epochs provided a valuable method for capturing wide variety of intensities intensity, although the epoch length may not fully account for the most intense activities, such as jumps. Nonetheless, they offer sufficient representation, and compared to 1-second epochs, they avoid capturing brief impacts that do not reflect actual activity patterns. Significant changes in detecting higher intensities are observed with epochs of 10 seconds or longer [38,39].

This study was not specifically designed to investigate why some individuals utilize their capacity while others do not. However, previous research suggests that walking is one of the most popular forms of physical activity among older adults [40]. Brisk walking and other hobbies including high-intensity activities (ball games, jogging, aerobic exercise), may help explain why participants in the high physical activity (PA) profiles accumulated more activity closer to their capacity compared to those in the low PA profiles. It can therefore be assumed that sports activities play a significant role in explaining why some people consistently engage in activities that approach in terms of intensity their physical capacity. However, this is undoubtedly related to psychological and environmental factors and personal preferences [41– 44]. Investigating these factors in the future is crucial for targeting interventions to increase physical activity. Additionally, based on this study, it can be inferred that even if physical capacity is already reduced, it is important to approach it habitually in order to maintain or at least slow down the decline in physical function. Obviously, safety should be considered to prevent adverse events, such as falls, or other medical emergencies potentially associated with vigorous activity.

When evaluating the results of the study, several limitations and strengths need to be considered. In this study, PC-PA profiles were constructed based on previously proposed categorization methods using a data-driven approach. A weakness of this approach is the potential loss of information due to grouping, as grouping inherently involves some loss of detail. However, characterizing the data within PC-PA classes can offer useful insights for more tailored and individualized physical activity promotion within this population. This study aims to address the well-known limitation that accelerometers cannot detect activities like swimming, cycling, carrying a child, walking uphill, or carrying a load [45] by examining the intensity distribution across all 5-second MAD bouts (Figure 2), which provides a broader understanding of activity patterns. The strength of the study lies in sufficient sample size of independently living older adults and its longitudinal design. Furthermore, the study benefits from the continuous device-based measurement of physical behaviour for multiple days [46,47] and employing a like-for-like capacity and free-living physical behaviour assessment.

## CONCLUSIONS

The findings suggest that engaging in PA close to one’s PC may help to protect against decline in physical functioning in older adults. Therefore, older adults should be encouraged to participate in physically demanding activities that could potentially maintain physical function.

## Acknowledgements

We thank the whole AGNES research team, especially Eeva-Maija Palonen and Niina Kajan and, all research assistants for their work in the data collection and analyses and all participants of the AGNES study for their time and effort. The authors wish to acknowledge CSC—IT Center for Science, Finland, for computational re-sources. The Gerontology Research Center is a joint effort between the University of Jyväskylä and the University of Tampere. The content of this article does not reflect the official opinion of the European Union. Responsibility for the information and views expressed in the article lies entirely with the author(s).

## Author Contributions

Conceptualization, A.L., K.L., L.P., E.P., T.R., Ti.R., L.K.; methodology, A.L., K.L., L.P., E.P., T.R., Ti.R., L.K.; formal analysis, A.L. and Ti.R.; writing—original draft preparation, A.L, K.L.; writing—review and editing, A.L., L.K., L.P., E.P., T.R., C.D., E.V.R. and Ti.R.; supervision, C.D., E.V.R. and Ti.R., L.K.; All authors have read and agreed to the published version of the manuscript.

## Data Availability Statement

After completion of the study, data will be stored at the Finnish Social Science Data Archive without potential identifiers (open access). Until then, pseudonymized datasets are available to external collaborators subject to agreement on the terms of data use and publication of results. To request the data, please contact Professor Taina Rantanen.

## REFERENCES

1. Landi F, Calvani R, Tosato M, Martone AM, Fusco D, Sisto A, et al. Age-Related Variations of Muscle Mass, Strength, and Physical Performance in Community-Dwellers: Results From the Milan EXPO Survey. Int J Environ Res Public Health.2021;18:88.e17-88.e24.

2. Valenzuela PL, Saco-Ledo G, Morales JS, Gallardo-Gómez D, Morales-Palomo F, López-Ortiz S, et al. Effects of physical exercise on physical function in older adults in residential care: a systematic review and network meta-analysis of randomised controlled trials. The Lancet Healthy Longevity.2023;4:e247–56.

3. Koeneman MA, Verheijden MW, Chinapaw MJM, Hopman-Rock M. Determinants of physical activity and exercise in healthy older adults: A systematic review. International Journal of Behavioral Nutrition and Physical Activity.2011;8:142.

4. D’Amore C, Saunders S, Bhatnagar N, Griffith LE, Richardson J, Beauchamp MK. Determinants of physical activity in community-dwelling older adults: an umbrella review. International Journal of Behavioral Nutrition and Physical Activity.2023;20:135.

5. Richards EA, Christ SL, Rietdyk S, Teas E, Franks MM. Association of Physical Activity and Gait Speed: Does Context Matter? American Journal of Lifestyle Medicine.2023;15598276231157311.

6. Leblanc A, Pescatello LS, Taylor BA, Capizzi JA, Clarkson PM, Michael White C, et al. Relationships between physical activity and muscular strength among healthy adults across the lifespan. Springerplus.2015;4:557.

7. Rostron ZP, Green RA, Kingsley M, Zacharias A. Associations Between Measures of Physical Activity and Muscle Size and Strength: A Systematic Review. Arch Rehabil Res Clin Transl.2021;3:100124.

8. Reijnierse EM, Geelen SJG, van der Schaaf M, Visser B, Wüst RCI, Pijnappels M, et al. Towards a core-set of mobility measures in ageing research: The need to define mobility and its constructs. BMC Geriatrics.2023;23:220.

9. Maetzler W, Correia Guedes L, Emmert KN, Kudelka J, Hildesheim HL, Paulides E, et al. Fatigue-Related Changes of Daily Function: Most Promising Measures for the Digital Age. Digit Biomark.2024;8:30–9.

10. Van Ancum JM, van Schooten KS, Jonkman NH, Huijben B, van Lummel RC, Meskers CGM, et al. Gait speed assessed by a 4-m walk test is not representative of daily-life gait speed in community-dwelling adults. Maturitas.2019;121:28–34.

11. van Lummel RC, Walgaard S, Pijnappels M, Elders PJM, Garcia-Aymerich J, van Dieën JH, et al. Physical Performance and Physical Activity in Older Adults: Associated but Separate Domains of Physical Function in Old Age. PLoS One.2015;10:e0144048.

12. Sievänen H. Impact loading—nature’s way to strengthen bone. Nat Rev Endocrinol.2012;8:391–3.

13. Koolen EH, van Hees HW, van Lummel RC, Dekhuijzen R, Djamin RS, Spruit MA, et al. “Can do” versus “do do”: A Novel Concept to Better Understand Physical Functioning in Patients with Chronic Obstructive Pulmonary Disease. J Clin Med.2019;8:340.

14. Orme MW, Lloyd-Evans PHI, Jayamaha AR, Katagira W, Kirenga B, Pina I, et al. A Case for Unifying Accelerometry-Derived Movement Behaviors and Tests of Exercise Capacity for the Assessment of Relative Physical Activity Intensity. Journal of Physical Activity and Health.2023;20:303–10.

15. Adams M, Carrascosa L, Jansen C-P, Ritter Y, Schwenk M. “Can Do” vs. “Do Do” in Older Adults: A Cross-Sectional Analysis of Sensor-Derived Physical Activity Patterns. Sensors.2023;23:1879.

16. Vähä-Ypyä H, Vasankari T, Husu P, Suni J, Sievänen H. A universal, accurate intensity-based classification of different physical activities using raw data of accelerometer. Clin Physiol Funct Imaging.2015;35:64–70.

17. Bull FC, Al-Ansari SS, Biddle S, Borodulin K, Buman MP, Cardon G, et al. World Health Organization 2020 guidelines on physical activity and sedentary behaviour. Br J Sports Med.2020;54:1451–62.

18. Sakari-Rantala R, Era P, Rantanen T, Heikkinen E. Associations of sensory-motor functions with poor mobility in 75- and 80-year-old people. Scand J Rehabil Med.1998;30:121–7.

19. Rantanen T, Pynnönen K, Saajanaho M, Siltanen S, Karavirta L, Kokko K, et al. Individualized counselling for active aging: protocol of a single-blinded, randomized controlled trial among older people (the AGNES intervention study). BMC Geriatr.2019;19:5.

20. Vähä-Ypyä H, Vasankari T, Husu P, Mänttäri A, Vuorimaa T, Suni J, et al. Validation of Cut-Points for Evaluating the Intensity of Physical Activity with Accelerometry-Based Mean Amplitude Deviation (MAD). PLoS One.2015;10:e0134813.

21. Perera S, Mody S, Woodman R, Studenski S. Meaningful Change and Responsiveness in Common Physical Performance Measures in Older Adults. Journal of the American Geriatrics Society.2006;54:743–9.

22. Bohannon RW. Comfortable and maximum walking speed of adults aged 20-79 years: reference values and determinants. Age Ageing.1997;26:15–9.

23. Guralnik JM, Ferrucci L, Pieper CF, Leveille SG, Markides KS, Ostir GV, et al. Lower Extremity Function and Subsequent Disability: Consistency Across Studies, Predictive Models, and Value of Gait Speed Alone Compared With the Short Physical Performance Battery. The Journals of Gerontology Series A: Biological Sciences and Medical Sciences.2000;55:M221–31.

24. Akobeng AK. Understanding diagnostic tests 3: Receiver operating characteristic curves. Acta Paediatr.2007;96:644–7.

25. Wang DXM, Yao J, Zirek Y, Reijnierse EM, Maier AB. Type and intensity distribution of structured and incidental lifestyle physical activity of students and office workers: a retrospective content analysis. Journal of Cachexia, Sarcopenia and Muscle.2022;11:3–25.

26. Guralnik JM, Simonsick EM, Ferrucci L, Glynn RJ, Berkman LF, Blazer DG, et al. A Short Physical Performance Battery Assessing Lower Extremity Function: Association With Self-Reported Disability and Prediction of Mortality and Nursing Home Admission. Journal of Gerontology.1994;49:M85–94.

27. Guralnik JM, Ferrucci L, Simonsick EM, Salive ME, Wallace RB. Lower-Extremity Function in Persons over the Age of 70 Years as a Predictor of Subsequent Disability. N Engl J Med.1995;332:556–62.

28. Folstein MF, Folstein SE, McHugh PR. “Mini-mental state”. A practical method for grading the cognitive state of patients for the clinician. Journal of Psychiatric Research.1975;12:189–98.

29. Mänty M, Heinonen A, Leinonen R, Törmäkangas T, Sakari-Rantala R, Hirvensalo M, et al. Construct and predictive validity of a self-reported measure of preclinical mobility limitation. Arch Phys Med Rehabil.2007;88:1108–13.

30. Rantakokko M, Portegijs E, Viljanen A, Iwarsson S, Rantanen T. Task Modifications in Walking Postpone Decline in Life-Space Mobility Among Community-Dwelling Older People: A 2-year Follow-up Study. J Gerontol A Biol Sci Med Sci.2017;72:1252–6.

31. Heikkinen E, Kauppinen M, Rantanen T, Leinonen R, Lyyra T-M, Suutama T, et al. Cohort differences in health, functioning and physical activity in the young-old Finnish population. Aging Clin Exp Res.2011;23:126–34.

32. Rantanen T, Avela J. Leg Extension Power and Walking Speed in Very Old People Living Independently. The Journals of Gerontology: Series A. 1997;52A:M225–31.

33. Rantanen T, Guralnik JM, Foley D, Masaki K, Leveille S, Curb JD, et al. Midlife Hand Grip Strength as a Predictor of Old Age Disability. JAMA.1999;281:558–60.

34. Liang K-Y, Zeger SL. Longitudinal data analysis using generalized linear models. Biometrika.1986;73:13–22.

35. SPSS. IBM Corp. released 2021. IBM SPSS Statistics for Windows, version 28.0. Armonk, NY: IBM Corp.2021;

36. R Core Team. R: A language and environment for statistical computing. R Foundation for Statistical Computing, Vienna, Austria. URL https://www.R-project.org/.2021; Available from: https://www.R-project.org/

37. Maula A, LaFond N, Orton E, Iliffe S, Audsley S, Vedhara K, et al. Use it or lose it: a qualitative study of the maintenance of physical activity in older adults. BMC Geriatr.2019;19:349.

38. Burchartz A, Kolb S, Klos L, Schmidt SCE, von Haaren-Mack B, Niessner C, et al. How specific combinations of epoch length, non-wear time and cut-points influence physical activity. Ger J Exerc Sport Res.2024;54:169–78.

39. Aibar A, Chanal J. Physical Education: The Effect of Epoch Lengths on Children’s Physical Activity in a Structured Context. PLoS One.2015;10:e0121238.

40. Chodzko-Zajko WJ, Proctor DN, Fiatarone Singh MA, Minson CT, Nigg CR, Salem GJ, et al. Exercise and Physical Activity for Older Adults. Medicine & Science in Sports & Exercise.2009;41:1510.

41. Kaspar R, Oswald F, Wahl H-W, Voss E, Wettstein M. Daily Mood and Out-of-Home Mobility in Older Adults: Does Cognitive Impairment Matter? J Appl Gerontol.2015;34:26– 47.

42. Feltz DL, Payment CA. Self-Efficacy Beliefs Related to Movement and Mobility. Quest.2005;57:24–36.

43. Hirvensalo M, Sakari-Rantala R, Kallinen M, Leinonen R, Lintunen T, Rantanen T. Underlying Factors in the Association between Depressed Mood and Mobility Limitation in Older People. Gerontology.2007;53:173–8.

44. Klenk J, Büchele G, Rapp K, Franke S, Peter R, the ActiFE Study Group. Walking on sunshine: effect of weather conditions on physical activity in older people:Figure 1. J Epidemiol Community Health.2012;66:474–6.

45. Lee I-M, Shiroma EJ. Using accelerometers to measure physical activity in large-scale epidemiological studies: issues and challenges. Br J Sports Med.2014;48:197–201.

46. Löppönen A, Karavirta L, Portegijs E, Koivunen K, Rantanen T, Finni T, et al. Day-to-Day Variability and Year-to-Year Reproducibility of Accelerometer-Measured Free-Living Sit-to-Stand Transitions Volume and Intensity among Community-Dwelling Older Adults. Sensors.2021;21:6068.

47. Pedersen ESL, Danquah IH, Petersen CB, Tolstrup JS. Intra-individual variability in day-to-day and month-to-month measurements of physical activity and sedentary behaviour at work and in leisure-time among Danish adults. BMC Public Health.2016;16:1222.

