## Supplementary File 1 for "Use it or lose it: A four-year follow-up to assess whether engagement with physical activity close to one’s physical capacity may protect against decline in physical functioning among older adults"

Supplementary File 1
Löppönen et al. 2024

According to the best knowledge of the authors, there are no thresholds available in the research literature for the MAD value of maximum 10-meter walking for this age group (75-85 years), based on which capacity can be classified as high or low. Therefore, in this study, the thresholds for 10-meter walking speed were determined separately for men and women. The determination was based on the Short Physical Performance Battery with cutoffs of 11-12 high and below 10 points low (Guralnik et al. 2000).

Receiver Operating Characteristics (ROC) analysis was performed to determine thresholds for 10-meter walking speed (Akobeng, 2007). The cut-points that best balanced the high sensitivity and high specificity of the test were calculated by finding the minimal value by using equation $(1 - \text{sensitivity})^2 + (1 - \text{specificity})^2$. The suitability of the test was evaluated by estimating the area under the curve (AUC). This value serves as a single measure that indicates the accuracy of the test: AUC 0.5 - 0.7 = low accuracy, AUC 0.7 - 0.9 = moderate accuracy, AUC > 0.9 = high accuracy (Akobeng, 2007).

For men, a cut-point of 0.73 G was determined with moderate accuracy (specificity 70%, sensitivity 66%, AUC 0.76), and similarly for women, the cut-point is 0.59 G (specificity 68%, sensitivity 76%, AUC 0.73) (Figure 1).

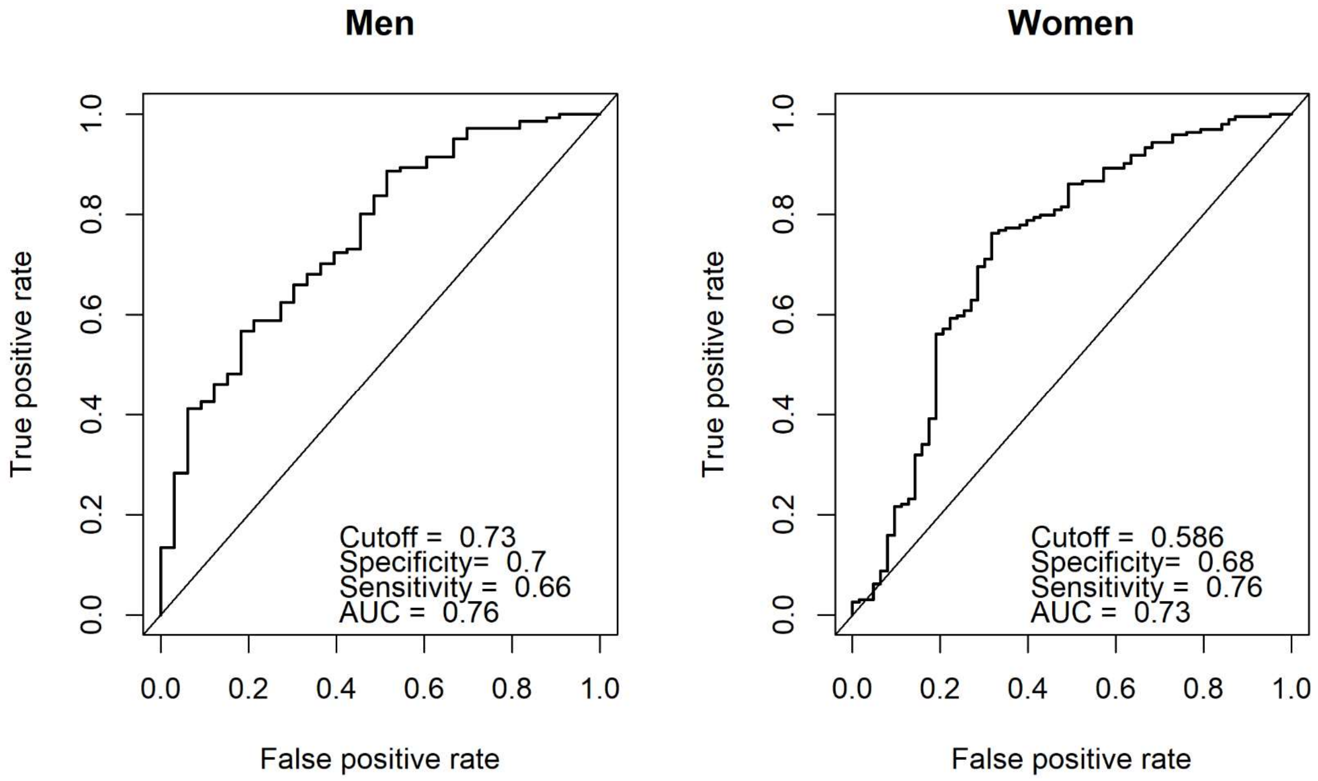


**Figure 1.**

**References**

Guralnik JM, Ferrucci L, Pieper CF, Leveille SG, Markides KS, Ostir GV, Studenski S, Berkman LF, Wallace RB. Lower extremity function and subsequent disability: consistency across studies, predictive models, and value of gait speed alone compared with the short physical performance battery. J Gerontol A Biol Sci Med Sci. 2000 Apr;55(4):M221-31. doi: 10.1093/gerona/55.4.m221. PMID: 10811152.

Akobeng, A. K. (2007). Understanding diagnostic tests 3: Receiver operating characteristic 3 curves. Acta Paediatrica, 96(5), 644-647
